# Epidemicity indices and reproduction numbers from infectious disease data in connected human populations

**DOI:** 10.1101/2024.02.22.24303216

**Authors:** Cristiano Trevisin, Lorenzo Mari, Marino Gatto, Andrea Rinaldo

## Abstract

We focus on distinctive data-driven measures of the fate of ongoing epidemics. The relevance of our pursuit is suggested by recent results proving that the short-term temporal evolution of infection spread is described by an epidemicity index related to the maximum instantaneous growth rate of new infections, echoing concepts and tools developed to study the reactivity of ecosystems. Suitable epidemicity indices can showcase the dynamics of infections, together with commonly employed effective reproduction numbers, especially when the latter assume values less than 1. In particular, epidemicity evaluates the short-term reactivity to perturbations of a disease-free equilibrium. Here, we show that sufficient epidemicity thresholds to prevent transient epidemic outbreaks in a spatially connected setting can be estimated by generalizing existing analogues derived when spatial effects are neglected. We specifically account for the discrete nature, in both space and time, of surveillance data of the type typically employed to estimate effective reproduction numbers that formed the bulk of the communication of the state of the COVID-19 pandemic and its controls. After analyzing the effects of spatial heterogeneity on the considered prognostic indicators, we perform a short- and long-term analysis on the COVID-19 pandemic in Italy, showing that endemic conditions were maintained throughout the duration of our simulation despite stringent control measures. Our method provides a portfolio of prognostic indices that are essential to pinpoint the ongoing pandemic in both a qualitative and quantitative manner, as our results demonstrate. We base our conclusions on extended investigations of the effects of spatial fragmentation of communities of different sizes owing to connectivity by human mobility and contact scenarios, within real geographic contexts and synthetic setups designed to test our framework.

**Author summary:** We revisit current standards in the characterization of the instantaneous state of epidemic spread in the light of newly acquired capabilities relaxing the assumption of spatially implicit disease geographies. Specifically, we generalize data-driven estimators of short-term epidemicity to include remotely acquired and objectively manipulated information about human mobility and the ensuing contacts between infected and susceptible individuals. Our results show that a spatial perspective can provide useful information on each node’s contribution to unfolding epidemics in connected systems. The level of heterogeneity that implies major departures among the key indicators, in particular epidemicity indices measuring the maximum growth rates of new infections (whose spatially explicit version is derived here), is identifiable and not unrealistic. Therefore, we suggest that only a portfolio of indicators based on epidemiological data (spatially implicit and explicit effective reproduction numbers and epidemicity indices that address the reactivity of the system) can provide a general, time-varying, and comprehensive overview of the effectiveness of epidemic control.

## Introduction

Contemporary epidemiological practice uses effective reproduction numbers (RNs), the estimate of the average number of secondary infections caused by a single primary infector, to compare alternative control measures [1–4]. To do so, the current standard is to employ reported infection data, often regardless of their spatial nature [5–16]. Effective RNs are commonly used as a proxy to gauge the spread of a pathogen in a population where values larger than unity indicate rampaging infections, while subunit values typically denote a receding epidemic and the asymptotic stability of the disease-free equilibrium (DFE) [17]. Containment measures (protective equipment, social distancing, vaccination campaigns, etc.) are usually tuned to guarantee that the effective RN stays below unity, effectively curbing the long-term course of the epidemic [18, 19]. Recently [20], a technological innovation made it possible to include additional data in the evaluation of spatially explicit RNs, typically big human mobility data gathered from cell phones and their GPS trackers. On the other hand, differences between spatially implicit and explicit RN estimators using surveillance data tend to diverge only if strong spatial heterogeneities exist in the disease transmission dynamics. Thus, current spatially implicit standards may prove reasonably accurate in connected and homogeneously populated contexts, but must be updated when these conditions are not met [20].

However, recent results suggest some caution in the use of RNs [21, 22]. In particular, the common practice of relaxing epidemiological containment measures when the effective RN decreases below 1 has been shown to potentially fail to achieve short-term control objectives [23]. By using reactivity analysis [24, 25], sufficient conditions were found that warrant a drastic curbing of local transient epidemic flares by preventing outbreaks due to imported cases [26, 27]. The question then arises as to whether these sufficient conditions can be achieved by suitably lowering the RN below a threshold *<* 1. It has been shown, within a spatially implicit context, that this is indeed possible [28]. As an example, in the case of COVID-19, safe RN thresholds should not exceed values in the range 0.24 ≤ RN ≤ 0.63, well below unit values, depending on certain theoretical assumptions [23, 28]. This may imply a rethinking of epidemiological practice.

What allows one to investigate the actual short-term temporal evolution of an epidemic is the definition of a suitable discrete epidemicity index [27], which is related to the maximum instantaneous growth of the infectious pool. As such, it can correctly capture the transient behaviour of the underlying epidemiological system. It should be noted, however, that in many of the existing theoretical approaches [26, 27], the epidemic spread was treated as a continuous process. While this assumption allows a realistic description of transmission dynamics, it may not fare well with the essentially discrete nature, in both space and time, of the surveillance data that are typically used to monitor the course of an ongoing epidemic. Furthermore, the estimate of sufficient conditions to prevent transient outbreaks was addressed in a spatially implicit context, an assumption that we relax in this work following previous attempts at estimating epidemicity in spatial settings [23, 29, 30]. Here, we utilize both a discrete-time and a space-explicit modeling approach. In this context, similarly to RN inference, an epidemicity index above unit suggests a temporary increase in the abundance of infectious individuals, while a sub-unit value ensures rapidly fading outbreaks that cannot spatially coalesce in larger flare-ups [23].

The computation of the discrete epidemicity index is of particular importance when the RNs take on values that are less than one because a system displaying long-term disease-free conditions may still develop a net increase in the number or prevalence of infectious (and therefore infected) individuals in the short term, which may lead to public health consequences whose severity depends on the size of the transient outbreak. Here, we show that the effective RN can be evaluated through so-called renewal equations in time and space, which connect the current generation of secondary infections (i.e., newly detected cases) to the active cases that became infected at some earlier time. This task was traditionally performed through a variety of methods that considered either spatial connectivity [20] or proxies thereof [4, 7, 10, 12, 14]. To compute the epidemicity index in a discrete-time setting, one needs to build a Leslie projection matrix [28]. Similarly to the related next-generation matrix method [1, 31–33], the Leslie matrix can be made time-dependent and updated to reflect the implementation of progressive policy measures. Moreover, it can also be made spatially explicit, which allows us to project the short-term dynamics of epidemic spread not only in time but also over space.

## Materials and methods

### A discrete infection process

Consider a metapopulation composed of *N* communities connected by human mobility. To this end, let the numbers in the infected compartment within a generic community *j* be defined as *I*_*j*_(*t, τ*), a function of both the current time *t* and the age of infection *τ* [32, 34, 35]. *Also, let C*_*lj*_(*t*) be the proportion of residents of community *j* who commute daily to community *l* for whatever purpose [20]. The governing equations in continuous time give rise to the following mathematical formulation [20, 28, 34]:

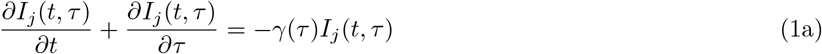

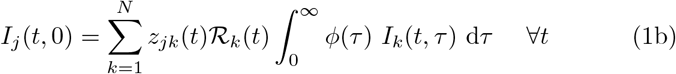

where: *γ*(*τ*) represents the instantaneous rate of exit from the infected compartment (because of recovery, death, quarantine, or isolation); *ϕ*(*τ*) is the rate of secondary transmission per single infectious case (per unity of RN); ℛ_*k*_(*t*) is the effective RN of community *k* at time *t*, namely, the number of secondary cases produced by one infectious case of community *k* at time *t* throughout the duration of the infection; and

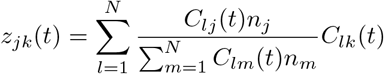

accounts for each node’s exposure to infectious individuals residing in a different community. The terms *z*_*jk*_(*t*) are computed to consider all possible means of contact and infection [20] by assuming homogeneous mixing of the population in the node where infection occurs. It should be noted that both **C** and **Z** are column-stochastic matrices, that is, Σ_*l*_ *C*_*lk*_ = Σ_*j*_ *z*_*jk*_ = 1. We define the probability of still being in the infected compartment at the age of infection *τ* as

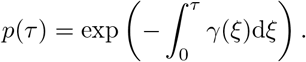

The probability density function of the generation times, *β*(*τ*), is related to this quantity via the equation

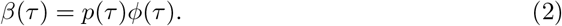

One may introduce a temporal discretization of the above problem so that it can be solved computationally with a suitable time step (e.g., one day), which is practical when epidemiological data (say, drawn from daily surveillance bulletins) are used to track the spatiotemporal evolution of an outbreak and/or produce epidemiological projections. The ensuing mathematical problem has already been addressed through the use of the method of characteristics [20]. Once this problem is tackled, a process discretized in both time and age of infection is obtained. It reads:

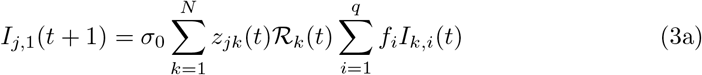

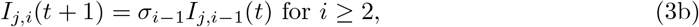

where: *I*_*j,i*_ is the number of infected people of infection age *i* (*i* = 1, …, *q*) in community *j* (*j* = 1, …, *N*), 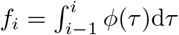 is a discretization of function *ϕ*(*τ*) over one day, *q* is the maximum age of infection, and

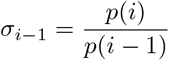

represents the proportion of infectious individuals with age of infection *i* − 1 that are still infectious on the next day.

### A spatially-explicit Leslie model

Let us now define a column vector **I**(*t*) that collects all the space-age of infection compartments at time *t*, that is:

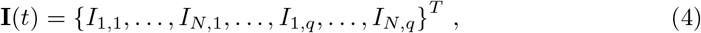

with *T* indicating matrix transposition. It is possible to rewrite equations (3) in matrix form as:

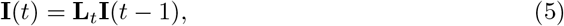

where matrix **L**_*t*_ is the Leslie projection matrix at time *t*. The elements of the Leslie projection matrix *L*_*j,k*_(*t*) [36] can be obtained by differentiating each equation by each state variable as follows:

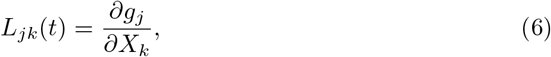

where *g*_*j*_ is the *j*-th equation shown in equation (3) and *X*_*k*_ is the *k*-th state variable (listed as in Eq. 4). For the process shown in model (3), the Leslie projection matrix is a block matrix that takes the following form:

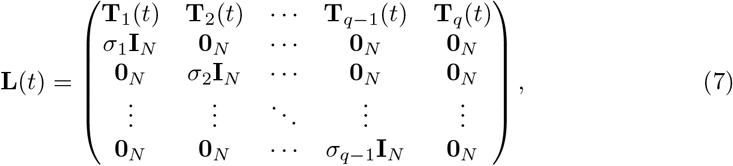

where **I**_*N*_ and **0**_*N*_ are the identity and null matrix of order *N*, respectively, and the submatrices **T**_*i*_(*t*) are defined as:

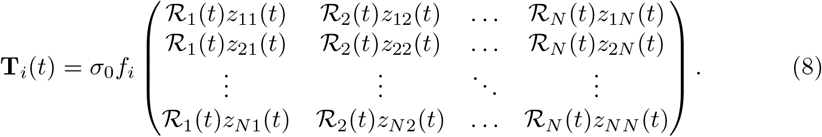

In order to carry out our reactivity analysis, we shall recur to two additional observed variables, the total abundance of infected people in a given node *j* at time *t* (**Y**_1_(*t*)) and their prevalence in the population (**Y**_2_(*t*)). The rationale for this choice is that, within real-time surveillance programs, the number of infected individuals is usually estimated as a whole and not for each age of infection. Also, decisions on intervention measures are usually taken at this level of data resolution. They are defined as:

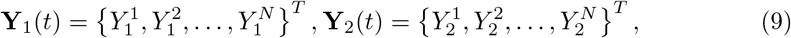

where:

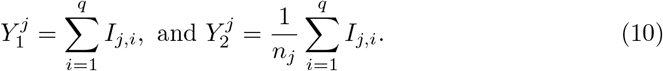

The above algebraic transformation can be done via the block matrices

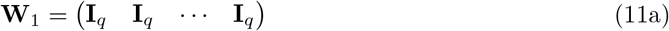

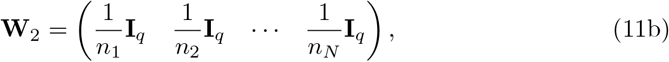

such that

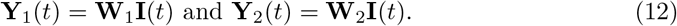

It should be noted that both **W**_1_ and **W**_2_ are full-rank matrices, that is, their nullspaces coincide with the zero vector. This implies that for any **I**(*t*) with at least one non-zero element, the corresponding output **Y**_*r*_(*t*) = **W**_*r*_**I**(*t*) (*r* = 1, 2) has at least one non-zero element. This property is a necessary condition to carry out our short-term reactivity analysis on the observed variables **Y**_1_(*t*) and **Y**_2_(*t*).

### Short-term reactivity analysis: the discrete epidemicity index and the amplification envelope

The evaluation of the Leslie projection matrix on a given day allows one to carry out a quantitative analysis of short-term epidemiological dynamics. Before continuing, let us briefly recall the computation of the algebraic *p*-norm (denoted as *ℓ*^*p*^) of a vector *x* of dimension *n*:

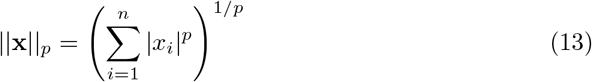

Let us now introduce the amplification envelope of a perturbation, which corresponds to the maximum growth of a trajectory departing from an equilibrium (in this case, the DFE) and is defined as:

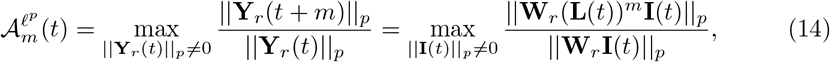

where *m* represents the number of days after which the perturbation’s growth is evaluated. Notable particular cases include the discrete epidemicity index, which is defined for *m* = 1 and corresponds to the maximum amplification of any perturbation in one time step:

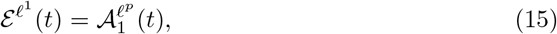

and the maximum amplification achieved by any perturbation at any time i.e.,

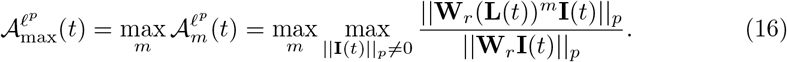

Note that the above formula is valid under the hypothesis that the Leslie projection matrix can be ‘frozen’ on day *t* and applied also in the following days, which is a reasonable assumption at least over relatively short time intervals.

Regardless of the epidemiological metric of interest, it is necessary to define which norm should be used to measure the amplifications. The most common choices are *ℓ*^1^ or *ℓ*^2^, where the former corresponds to the total absolute deviation of a trajectory originated by a perturbation of the DFE, while the latter refers to its Euclidean distance. In both cases, the epidemicity index can be formally (and in some cases analytically) defined. The choice of the norm addresses different information regarding the fate of a perturbation and has been shown to impact the results of the analysis [28]. In the following, we shall conduct our reactivity analysis using the output variable **Y**_1_ (total number of infectious individuals in each local population) as measured by the *ℓ*^1^-norm, as well as the output variable **Y**_2_ (prevalence of infectious individuals in each local population) as measured by both the *ℓ*^1^- and the the *ℓ*^2^-norm. Because the most populated nodes of a metapopulation are more likely to record more infectious individuals, the use of the *ℓ*^2^-norm with the output variable **Y**_1_ would overshadow the epidemiological role of other, less populated nodes, regardless of whether their healthcare infrastructure could potentially be under severe strain.

#### *ℓ*^1^**-norm**

Applying this norm to a vector **Y**(*t*) corresponds to evaluating the sum of the vector components at time *t*, that is:

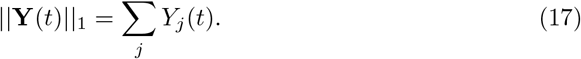

Therefore:

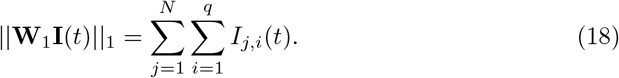

is the total abundance of infectious individuals in the whole metapopulation at time *t*. By using the multiplication by the Leslie projection matrix, and by recalling the column-stochasticity of matrix **Z**, one finds out that

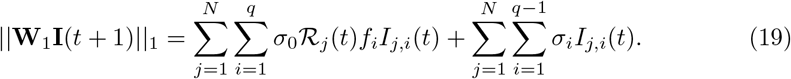

If we let *σ*_*q*_ = 0, the above expression reduces to:

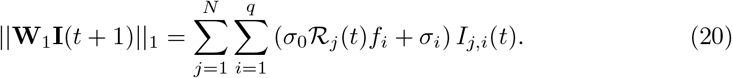

Therefore, the problem of finding the 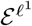 epidemicity is equivalent to:

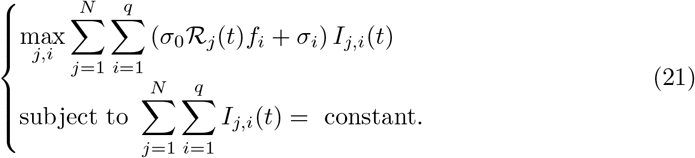

This is a linear programming (LP) problem in the variables *I*_*j,i*_(*t*). Since there is only one equality constraint, the optimal solution has just one basic variable, precisely the one characterized by max_*j,i*_ (*σ*_0_ ℛ _*j*_(*t*)*f*_*i*_ + *σ*_*i*_). For this well-known result, see e.g., Luenberger (1973) [37]. One can of course set the constant to one so that *I*_*j,i*_ = 1 and the epidemicity index reads:

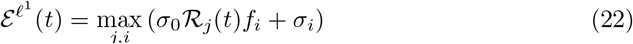

If we assume *σ*_*j*_ = *σ ∀j < q*, and *f*_*q*_ = 0, then:

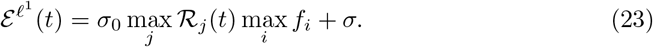

This result shows that the maximum one-step growth of the system depends on the maximum local effective RN. As such, whenever any local effective RN exceeds the threshold 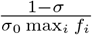 [28] the system is reactive, which implies that there exists at least one perturbation that develops a growing response associated with a transient epidemic outbreak.

The *ℓ*^1^-norm can be used for the prevalence of infectious people. In this case, the norm is proportional to the average prevalence across the different communities. As such, a decision maker might be interested in avoiding its increase over time. The relevant algebra becomes slightly more complex. Let us consider the norm

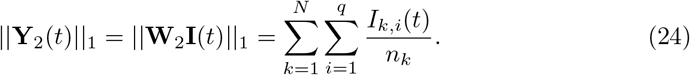

Multiplying by the Leslie matrix, at time *t* + 1 we have

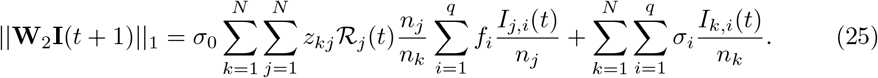

Let us introduce the 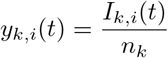, thus obtaining a revised form of the above formula, which now reads:

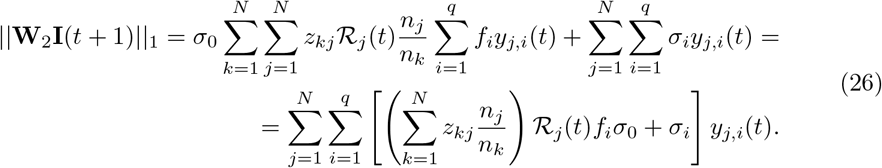

Therefore, the problem of finding the epidemicity index, that is 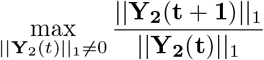 is equivalent to the following LP problem:

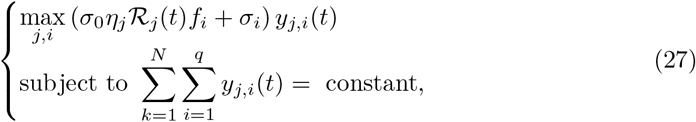

with 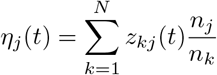. The solution has one basic variable, the one characterized by:max_*j,i*_ (*σ*_0_*η*_*j*_(*t*)ℛ_*j*_(*t*)*f*_*i*_ + *σ*_*i*_). The epidemicity index is given by

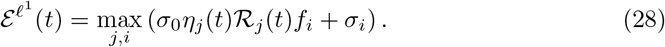

It should be noted that if all *n*_*k*_ are equal, then this solution would yield the one shown in Eq. (22). If we assume *σ*_*j*_ = *σ ∀j < q*, and *f*_*q*_ = 0, then:

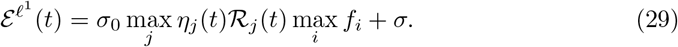

Therefore, if we consider the prevalence to establish epidemicity, the focal community might not be the one with the maximum RN, but the one with the maximum *η*_*j*_(*t*)ℛ_*j*_(*t*), which depends on the interplay between population size, connectivity, and local RNs.

When the *ℓ*^1^-norm is chosen, it may be of interest to consider the epidemicity subset, which is the ensemble of nodes that can produce transient responses [38]. The epidemic subset is defined as:

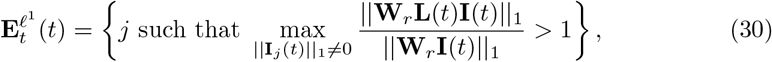

where we used **I**_*j*_(*t*) to indicate the set of infected people in node *j*. More simply, the elements of the epidemic subset are defined for the observed variable **Y**_1_(*t*) as the nodes corresponding to:

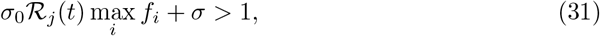

and, for the observed variable **Y**_2_(*t*) as:

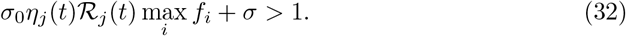

Any perturbation to the DFE acting upon one or more of these age-space combinations would thus be at least temporarily amplified, thereby developing a transient epidemic outbreak. As an example, these perturbations may be associated with the arrival of infected individuals with specific ages of infection in specific communities of the spatial network, provided that the local effective RNs allow for epidemic behaviour.

#### *ℓ*^2^**-norm**

In this case, the norm of the infectious compartment is defined as the Euclidean distance from the DFE, i.e.:

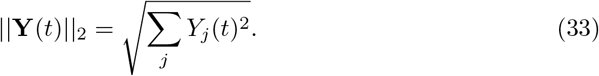

Thus, the *ℓ*^2^-norm accounts for the behaviour of the whole system, but emphasizes superlinearly the fastest growing local components of the perturbation. In order to evaluate the effect of mobility on the *ℓ*^2^-norm epidemicity index, we measure the divergence between the epidemicity indices computed with connected versus disconnected nodes. To this end, we resort to the mean absolute percentage deviation, which is defined as:

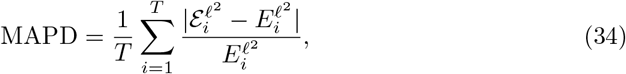

where 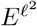 denotes the *ℓ*^2^-norm epidemicity index corresponding to a metapopulation with all *ξ*_*j*_ = 0 *∀j* (i.e., a metapopulation where the connectivity matrix **C** is an identity matrix).

### Long-term dynamics: the global reproduction number

A simple way to assess the long-term dynamics of an epidemic usually consists of evaluating the RN, which is commonly done via the next-generation matrix [1, 31–33]. While this approach has been extensively used for continuous-time models, it has not yet found many applications in discrete-time epidemiological systems [32].

Following the canonical next-generation matrix approach, let us decompose the Leslie projection matrix into two matrices, a transmission matrix **T**(*t*), containing all the terms reflecting the new cases (see Eq. (3)a), and a transition matrix **Σ**(*t*), containing the transition terms (see Eq. (3)b), such that **T**(*t*) + **Σ**(*t*) = **L**(*t*). The next-generation matrix can be built as [32, 39]:

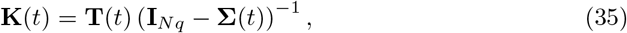

where **I**_*Nq*_ is the identity matrix of size *Nq × Nq*. The global RN of the system is defined as the spectral radius of the next-generation matrix:

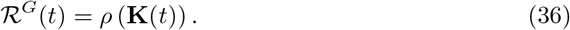

This quantity determines the long-term behaviour of the system as a whole and also bears consequences on the amplification envelope. In fact, if ℛ^*G*^(*t*) *>* 1 then the amplification envelope grows unbounded over time (i.e., 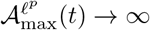 if *t* → ∞). This relationship implies that in a spatially connected system where local effective RNs are either above or below the unit threshold, whenever ℛ^*G*^(*t*) *>* 1 there exists at least one possible perturbation of the DFE that could be indefinitely amplified over time. In practice, however, the finite size of the population and the nonlinearity of the transmission process would prevent us from observing this behaviour.

### Assumptions on COVID-19

To calculate the distribution of generation times, we use the estimates reported in [40], where a mean generation time of 5.2 days and a standard deviation of 1.7 days are documented. We estimate a mean duration of infection (including incubation and a pre-symptomatic phase followed by either symptomatic or asymptomatic periods) equal to 11.7 days (which corresponds to a constant value *σ*_*j*_ = exp(−0.068) *∀j* ≥ 1) [41]. Finally, we assume *σ*_0_ = 1 for the sake of simplicity.

### Experiments on a synthetic metapopulation

In some of our experiments, we resort to a fictitious metapopulation of the kind used in [20]. Specifically, we assume that the metapopulation is composed of *N* connected local communities, whose population abundance is randomly generated from a Zipf distribution with an exponent equal to −2 and a minimum value generated from a uniform distribution with support between 10^4^ and 10^5^. The number of communities is also drawn from a discrete uniform distribution whose support includes all natural numbers in [2, 10]. The proportion of outgoing mobility *ξ*_*l*_ = Σ_*j≠l*_ *C*_*lj*_ is drawn from a normal distribution with reflecting boundaries (to ensure values between 0 and 1) and prescribed mean/standard deviation values. For mobile individuals, the probabilities of reaching different destinations are randomly generated from a uniform 0–1 distribution and normalized to *ξ*_*l*_. For the non-spatial case, it has been demonstrated that there is a clear relationship between the (effective) RN and the epidemicity index, and that this relationship can be estimated analytically [28]. The local effective-RNs are generated from a log-normal distribution with a mean of 1.05 and a standard deviation of 0.33.

### Experiments on a two-node synthetic metapopulation

In some cases, we use an even simpler, two-node synthetic metapopulation with local population abundances *n*_1_ and *n*_2_. We further define the local RNs as ℛ_1_ = 0.5, ℛ_2_ = 1.5. The mobility fluxes between the two nodes are described by the connection matrix

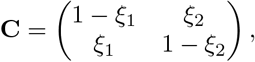

where *ξ*_1_ and *ξ*_2_ quantify the outgoing mobility from either of the two nodes.

### Experiments on the COVID-19 pandemic in Italy

In what follows, we will resort to the COVID-19 pandemic in Italy between January 24, 2020 and October 15, 2021 as a testbed for our analytical framework. The rationale behind this choice, in particular concerning the selected time window, is that suspected cases linked to the BA.1 (Omicron) variant, which substantially altered the epidemiological parameters (such as the distribution of generation times), were detected in Italy shortly after this period. Extending the temporal window of analysis would thus require a multi-variant approach, which lies outside the scope of this paper. We consider *N* = 20 local communities, corresponding to the regions of Italy (first-level administrative divisions). Epidemiological data is retrieved from the Italian Department of Public Protection (https://github.com/pcm-dpc/COVID-19). It should be noted that, for the sake of standardization, data from the autonomous provinces of Trento and Bolzano are merged in a single region Trentino-Sudtirol (while they correspond to different NUTS-2 entities for statistical purposes). To evaluate matrices **C** and **Z**, as well as the local effective RNs, we resort to the same technique implemented in [20], which is based on Eq. (3a), with *I*_*j*,1_ representing the new infections observed in node *j* at time *t*. Note that the RNs calculated in this way do not correspond to the global RN introduced above; rather, they represent sequences of regional-level RNs that also account for the local connectivity patterns of each region. The pre-pandemic connectivity matrix is estimated through mobility data obtained from the Italian Institute of Statistics (ISTAT) (accessible at: https://www.istat.it/it/archivio/139381) and updated during the COVID-19 pandemic through the “Workplace mobility” information provided by the Google Community Mobility Reports (https://www.google.com/covid19/mobility/). The computation of the aforementioned matrices is identical as in [20], except for a coarser spatial resolution (regional vs. provincial) being chosen to compensate a wider spatial domain (national vs. regional).

## Results

### Effect of mobility on the global reproduction number and epidemicity indices

We first tested the influence of the connectivity of the system on the value of the global effective RN, ℛ^*G*^(*t*), using a two-node synthetic metapopulation as described in the Materials and Methods section. Specifically, we investigated how ℛ^*G*^(*t*) relates to the largest local effective RN and to the weighted mean of the local effective RNs (where the weights are assumed equal to the proportion of the total resident population in the corresponding node). Our results suggest that

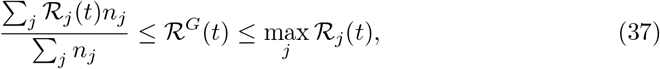

where *n*_*j*_ is the population of the *j*-th community, and ℛ_*j*_(*t*) is its local effective RN.

The first result we report (Fig 1) is that the global RN may take sub-unit values even is one node has a local RN above the unit threshold. Under such scenario, the number of expected infections does not grow indefinitely over time, but rather tends to zero asymptotically. The global RN is indeed defined as a function of not only the local RNs, but also the local populations and the mobility fluxes among the interconnected communities. We find that the global effective RN strongly depends on the differences in population size between the two nodes and the configuration of the outgoing fluxes (Fig 1). Indeed, for all the tested parameter combinations, the global RN appears to be maximum (i.e., equal to 1.5 in our numerical example) for either zero or full outgoing mobility (i.e., when the new infections in each node depend only on the active cases in another, unique node). For well-mixed mobility fluxes (both *ξ* equal to 0.5, where *ξ* represents the proportion of outgoing mobility from each node), the global effective RN takes its minimum value, which is equal to the population-weighted average of the local RNs. We find singularities when only one of the *ξ* is null. Because of the definition of the spatial exposure matrix **Z**, in such cases the active population in either of the two nodes is null, which prevents the computation of the epidemiological indices. Finally, a central symmetry holds for all combinations of the two mobility-related parameters. When different configurations of the local population sizes are tested, the lower bound of the global RN changes following the relationship shown in Eq. (37). A larger population in the node with the largest local RN results in higher values of the global RN across the tested mobility combinations, whereas the opposite induces a drastically lower minimum value of the global RN.

**Fig 1.**
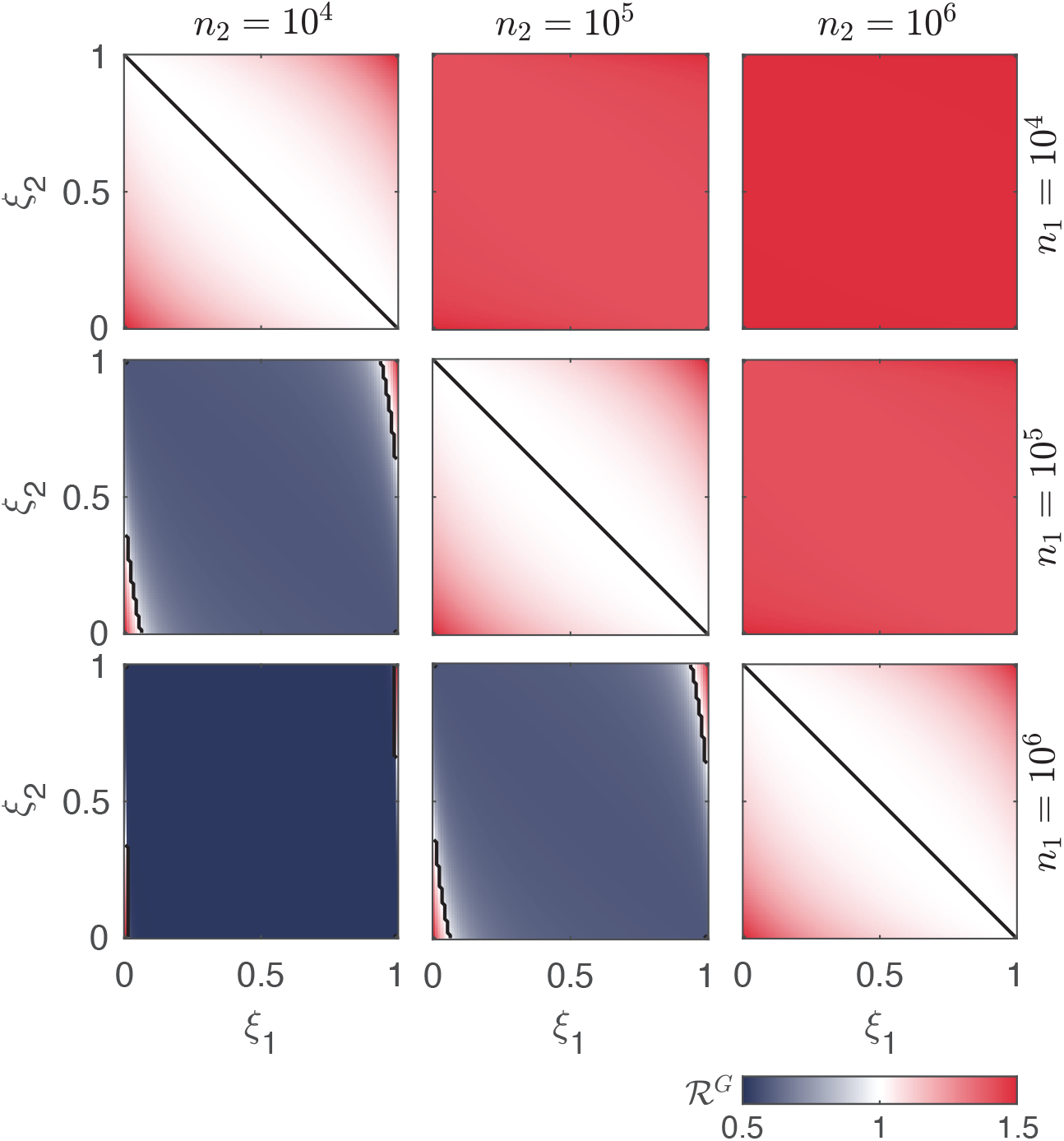
Global RN in a simplified network of two nodes as a function of the proportion of mobile individuals. The black lines correspond to the unit threshold of the global RN. Different population size combinations are tested in the different panels. The two nodes have local RNs ℛ_1_ = 0.5 and ℛ_2_ = 1.5. Mobility fluxes are defined through the coefficients of outgoing mobility from each node (*ξ*_1_ and *ξ*_2_; see Materials and Methods).

Moreover, we tested the impact of the two local RNs with fixed mobility fluxes (*ξ*_1_ = *ξ*_2_ = 0.25) for different configurations of the population distribution (Fig 2). We found that both local RNs and population sizes substantially impact the values of the considered epidemiological indices (highlighting whether the system may asymptotically converge to a stable DFE and, if so, whether this equilibrium is reactive). In a simple scenario with equal populations in the two nodes, the condition for the asymptotic stability of the DFE (*R*^*G*^ *<* 1, which applies when both local RNs are *<* 1 or when one is above the unit threshold and the other is sufficiently below it) does not automatically guarantee non-reactive conditions. Indeed, panel A of Fig 2 shows that additional thresholds for both local RNs must be met to guarantee that both the *ℓ*^1^- and the *ℓ*^2^-norm epidemicity indices take sub-threshold values, similarly to previous finding for continuous-time systems [23]. When the population is heterogeneously distributed (panels B–D), with one local population being substantially larger than the other, the global RN almost uniquely depends on the RN of the most populated node, with little dependence on the other local RNs. The global RN displays high sensitivity to the largest local RN whenever the *n*_2_*/n*_1_ ratio increases, while this behaviour is less pronounced for the considered epidemicity indices,as shown in Fig 2. However, when the population sizes are extremely different from each other (e.g., in the case of Fig 2D, where *n*_2_*/n*_1_ = 50), then the epidemicity indices computed on the prevalence of infectious individuals acquire an almost exclusive dependence on the RN corresponding to the largest local population. Finally, the threshold for the *ℓ*^1^-norm epidemicity never changes and is slightly larger than 0.2, which is consistent with previous spatially-implicit investigations [28].

**Fig 2.**
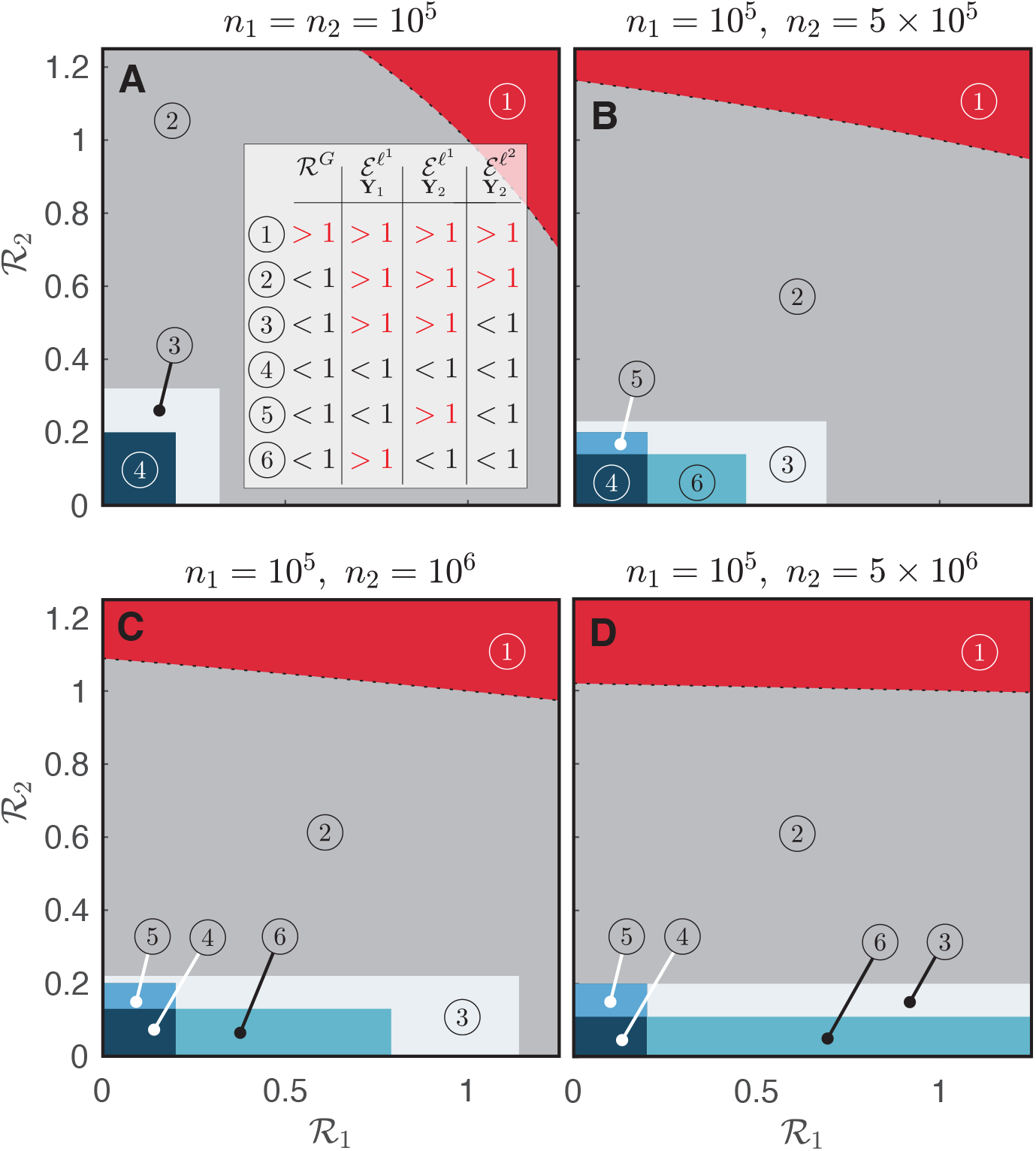
Comparative analysis of the epidemiological indices in a metapopulation with two nodes. Dependence on the two local reproduction numbers of the global reproduction number, the *ℓ*^1^-norm of both observed variables (**Y**_1_ and **Y**_2_, respectively the total number of infectious individuals and the prevalence of infectious individuals in each local population), and the *ℓ*^2^-norm epidemicity index (based on variable **Y**_2_). Colors code different configurations of the epidemiological indices, leading to different short- and long-term disease transmission scenarios (inset of panel **A**). Specifically, red denotes endemic conditions (1), dark grey represents epidemic but non-endemic conditions (2), light grey represents a scenario where the system is reactive in the *ℓ*^1^-norm applied to both output transformations but not in the *ℓ*^2^-norm applied to the **Y**_**2**_ transformation (3), and the blue hues represent non-reactivity in at least one of the two considered *ℓ*^1^-norms (4–6). For both nodes, *ξ*_1_ = *ξ*_2_ = 0.25.

We also stress that, for given combinations of the two local RNs, there exists a scenario where the *ℓ*^1^-norm epidemicity index is below the reactive threshold and its *ℓ*^2^ analogue is not. Under our output transformation matrices **W**_1_ and **W**_2_, this implies that the total number of infectious people is increasing, yet the *ℓ*^2^-norm of the local prevalences of infectious individuals is decreasing. Another interesting result concerns the comparison between the *ℓ*^1^-norm computed on the total number of infectious individuals (see Fig 2, where non-reactivity is represented by zones 4 and 5) and that related to their prevalence in each local population (zones 4 and 6 in Fig 2). The two observed variables do not share their reactivity thresholds unless all populations are equal, as shown in our analytical derivations (see Materials and Methods). Indeed, whenever one node has a population that is substantially larger than the other node’s, the threshold of the RN above which reactive conditions on the prevalence of infectious individuals apply decreases substantially. In our two-node example, it can be seen that this threshold is halved when the most populous node has a fifty-fold larger population than the other, as shown in Fig 2D. Similarly to the *ℓ*^2^-norm, we also remark that the reactivity of the output variable **Y**_2_ as measured with the *ℓ*^1^-norm is almost uniquely dependent on the RN of the node with the highest value when substantially different population sizes are involved, as shown by the elongated shapes of the regions marked as 6 in Fig 2.

We also measured how the *ℓ*^2^-norm epidemicity index changes when higher mobility fluxes (as determined by the coefficients of outgoing mobility *ξ*_*j*_ (*j* = 1, …, *N*)) depart from a baseline configuration without mobility. To this end, we resorted to a synthetic metapopulation of *N* communities with Zipf-distributed population sizes (Materials and Methods). We find (Fig 3, panels A–I) lower values of the *ℓ*^2^-norm epidemicity index for increasing values of the outgoing mobility parameter *ξ*, which is also reflected in the growth of the mean absolute percentage deviation (MAPD, Fig 3J) with respect to the scenario without mobility. We also note that, for larger outgoing mobility fluxes (*ξ >* 0.05), an increase in the number of the interconnected communities further amplifies the deviation from the no-mobility scenario (Fig 3K).

**Fig 3.**
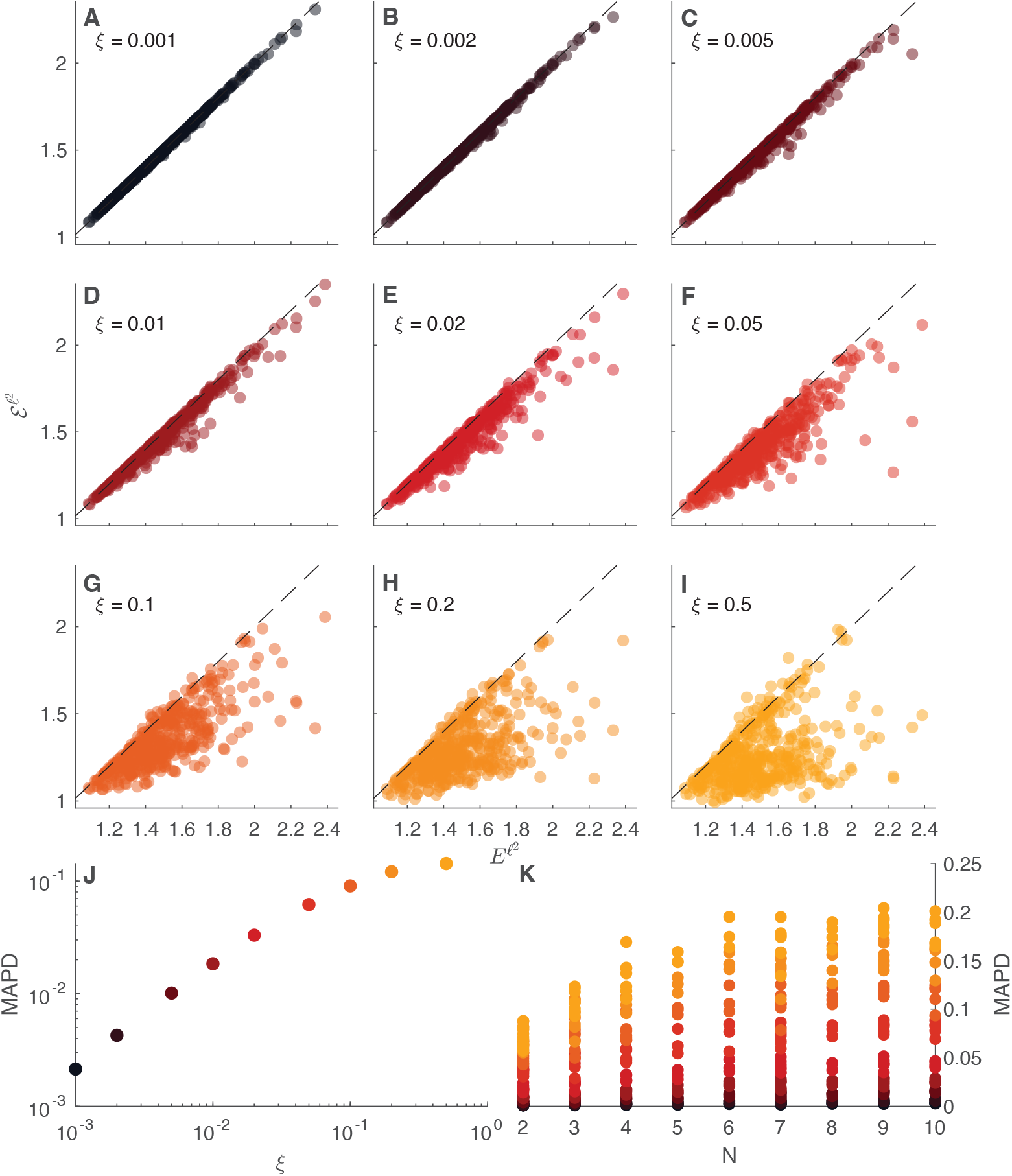
Impact of increasing outgoing mobility fluxes on the *ℓ*^2^-norm discrete epidemicity index based on the prevalence of infectious people. **A**-**I**: Scatter plots of the disconnected *ℓ*^2^-norm epidemicity index 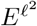 versus the spatially connected epidemicity index 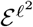 for different values of the outgoing mobility parameter *ξ*. The dashed black lines represent the identity relationship. **J**: MAPD between the values of the connected and disconnected *ℓ*^2^-norm epidemicity index, as a function of outgoing mobility *ξ*. **K**: As in the left-bottom panel, but MAPD is shown as a function of both outgoing mobility *ξ* (color coded) and the number of interconnected communities *N*.

### Computational experiments on the COVID-19 pandemic in Italy

In the following, we provide a practical example of the computation of the epidemicity indices in a realistic spatially explicit setting. To that end, we refer to the epidemiological data from Italy described in the Materials and Methods section, where details on the time-varying contact matrix and effective RNs can also be found. Our results are summarized in Fig 4. We observe that the global effective RN is quite close to the largest local effective RN for most of the considered period (panel A), owing to the fairly low mobility coefficients (the largest value reached by any *ξ*_*j*_ is equal to 0.045). The global effective RN, depending on whether the epidemic is waning or revamping, oscillates across the unit threshold, which leads to the maximum value of the amplification envelope (panels B and C) taking non-finite values at times. In contrast, the epidemicity indices always take larger-than-unit values, regardless of the chosen norm. Given our choice of transformation matrices **W**_1_ and **W**_2_, we find that the two epidemicity are very close (0.4% mean absolute percentage deviation), with the values computed with the *ℓ*^1^-norm being slightly larger than their *ℓ*^1^-norm counterparts. An analysis of the epidemic subset shows that, for both norms and output transformations, all regions were reactive during the whole timespan of our analysis. This finding suggests that suitable perturbations to the DFE (e.g., in the form of imports of cases with an age of infection close to the peak in transmissibility) could have produced transient (if *R*^*G*^(*t*) *<* 1) or sustained (if *R*^*G*^(*t*) *>* 1) epidemic outbreaks regardless of the region(s) where the perturbation was localized and the time when it occurred. We also compared (Fig 4B) the *ℓ*^1^-norm epidemicity indices based on the total number of infectious individuals and their prevalence within the local populations. We found that, although some differences can be spotted, they are not quantitatively large, with a mean absolute percentage deviation (with respect to **Y**_1_) of 0.56% for the *ℓ*^1^-norm epidemicity index and of 5.56% for the maximum value of the corresponding amplification envelope.

**Fig 4.**
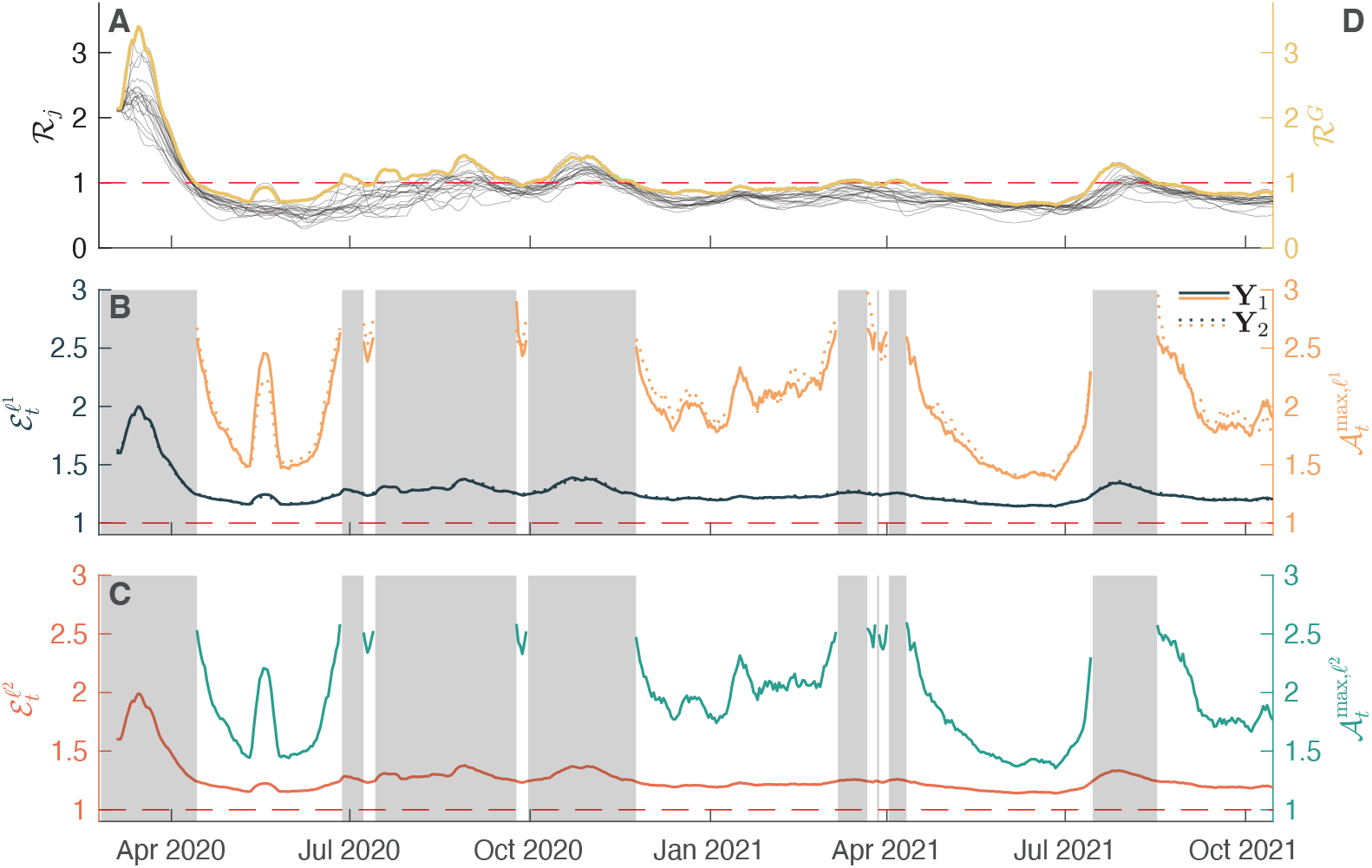
Reactivity analysis of the covid-19 epidemics in Italy. **A**: Local effective RNs inferred from each region of Italy (thin gray lines) and global effective RN (thick yellow line). **B**: *ℓ*^1^-norm epidemicity index (left axis, dark blue) and maximum value of the amplification envelope (right axis, yellow) of the variables **Y**_1_ (total number of infectious individuals in each community) and **Y**_2_ (prevalence of infectious individuals in each community). **C**: *ℓ*^2^-norm epidemicity index (left axis, red) and maximum value of the amplification envelope (right axis, green). In panels **B** and **C**, gray patches denote *R*^*G*^(*t*) *>* 1 (in which case the amplification envelope grows unbounded over time). The Bayesian method used to compute the local effective RNs [20] produces numerically close values of the local effective RNs at the beginning of the covid-19 epidemic. This reflects the fact that the epidemic wave does not affect all nodes at the same time.

A correlative analysis between each local effective RN, the global effective RN, and the different epidemicity indices has been carried out too. Our analysis shows that each local effective RN correlates in a similar way to each of the other considered epidemiological indices. We find that both the global effective RN and the largest local effective RN correlate almost perfectly with the *ℓ*^1^-norm epidemicity computed on output variable **Y**_1_, with a correlation coefficient of 0.9994 and 1, respectively. They also correlate with the *ℓ*^2^-norm epidemicity calculated on the prevalence of infectious individuals in the local populations (**Y**_2_), with coefficients of 1 and 0.9994, respectively. Because the covid-19 epidemic in Italy has often produced synchronous effects in many Italian regions, there also is high correlation between each local effective RN and both the global effective RN and the *ℓ*^2^-norm epidemicity index, with coefficients ranging between 0.84 (Molise) and 0.98 (Lazio). However, it can be seen that the fraction of time each region dominates from an epidemicity perspective (e.g., a perturbation in that node of the metapopulation would produce the largest short-term growth in the system output) is different for the two compared variables, owing to variations of the *η*_*j*_(*t*) coefficient (Materials and Methods). These differences in the share of time each node dominates across the two different output variables also reflect different epidemicity thresholds for the local RN to guarantee non-reactive conditions. While these variations in the threshold values are not particularly strong, especially given the homogeneity in mobility fluxes across the different Italian regions, they do exist. The region that requires the lowest RN threshold to warrant non-reactive conditions is Apulia (with a limit of 0.202) which is also the region with the highest share of time as the dominant node (Fig 5). The region that would require the least stringent threshold is Molise (whose limit for the RN is 0.226).

**Fig 5.**
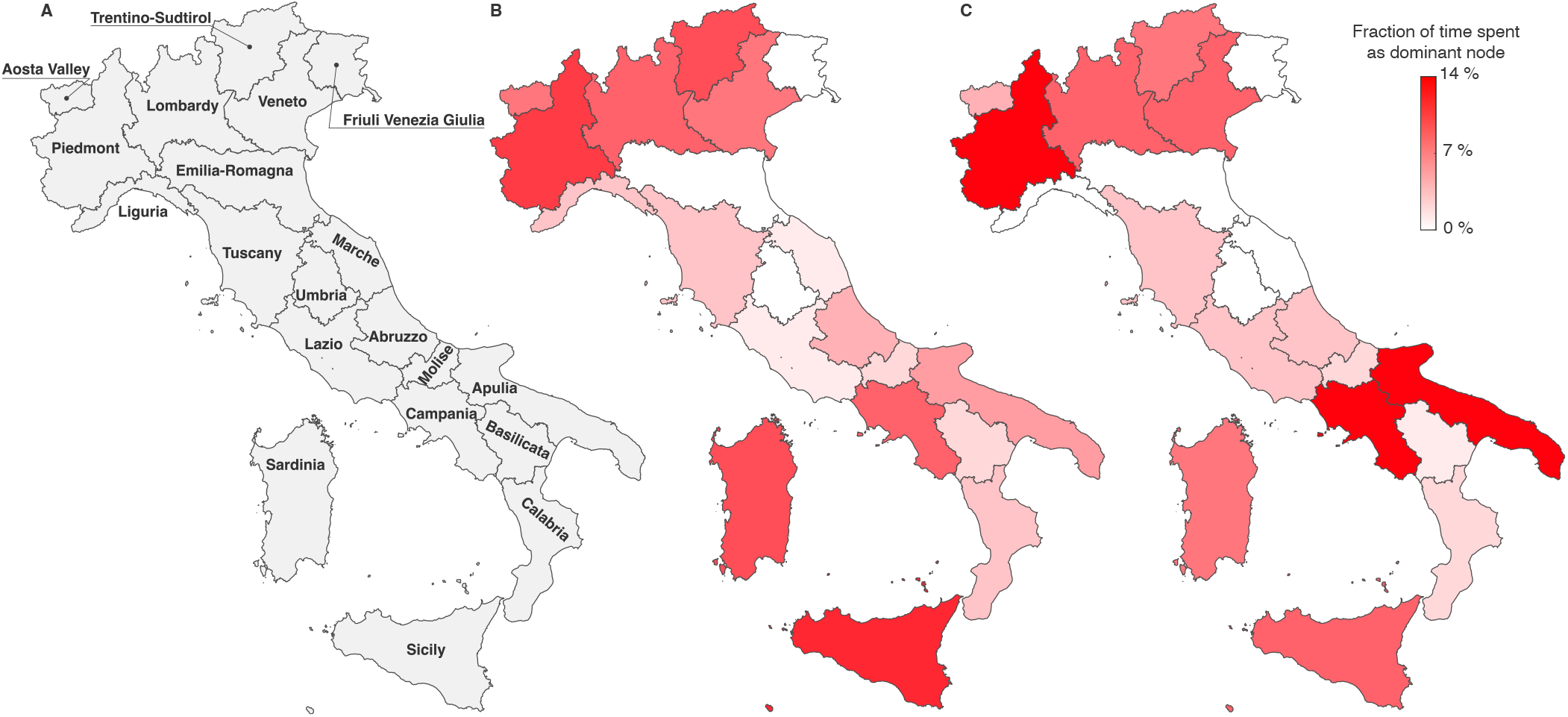
Share of the time each Italian region spends as dominant node from an epidemicity perspective. **A**: Map of Italy and its regions. Please note that, for graphical purposes, the position of Sardinia has been slightly displaced eastwards and some Mediterranean islands have been removed. **B**: Fraction of time spent as dominant node in the epidemic subset computed with the total number of infectious individuals in each region (observed variable **Y**_1_(*t*) with the *ℓ*^1^-norm). **C**: Fraction of time spent as dominant node computed on the prevalence of infectious individuals in each region (observed variable **Y**_2_(*t*) with the *ℓ*^1^-norm).

Finally, to disentangle the effect of each region’s local RN on the global RN and the *ℓ*^2^-norm epidemicity index, we carried out a sensitivity analysis where we added a *±*20% variation to each time series of local effective RNs (as shown in Fig 4A), one at a time. This analysis is shown in Fig 6. Positive 20% variations of one local RN often result in a positive 5 − 10% variation of the global RN and a positive 0.5 − 2% variation on the epidemicity indices, with similar effects on each index. We also find a negative correlation (with coefficient −0.90) between positive variations of the *ℓ*^1^-norm of the epidemicity index based on the prevalence of infectious individuals in each node and the nodal thresholds for the RN guaranteeing non-reactivity. On the other hand, only reducing one local effective RN at a time produces little to no impact on any of the considered epidemiological indices. This is also justified by all regions being always well above the reactivity threshold.

**Fig 6.**
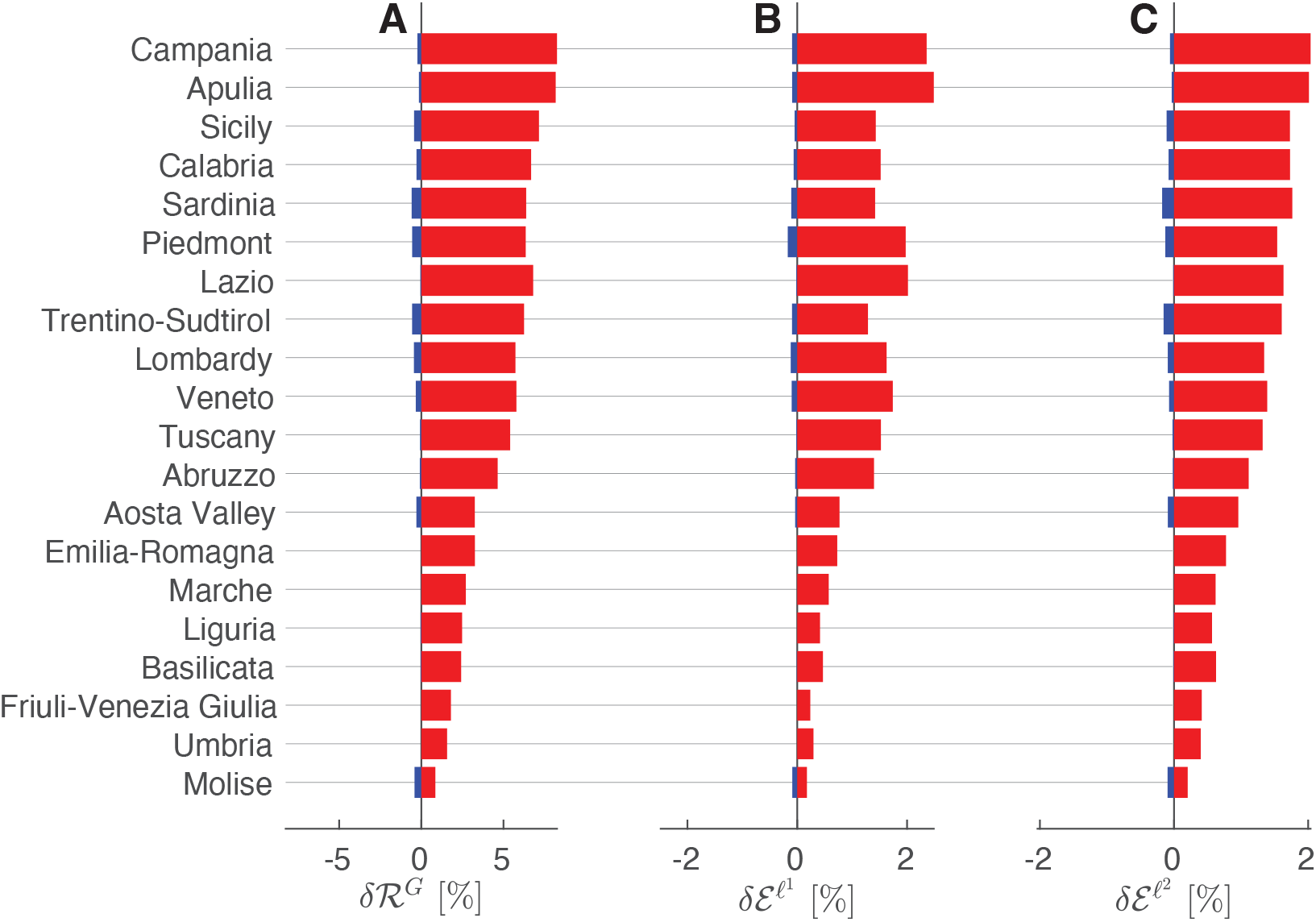
Sensitivity analysis of selected epidemiological indicators evaluated in the context of the covid-19 pandemic in Italy. Mean percentage deviation of the time series of the global reproduction number (panel **A**), and of the *ℓ*^1^- and *ℓ*^2^-norm epidemicity indices (panels **B** and **C**, respectively) of the prevalence of infected individuals (observed variable **Y**_2_), using a *±*20% variation of the RN time series in each region with respect to the baseline estimates shown in Fig 4. The effects of positive perturbations are shown in red, whereas negative perturbations are shown in blue.

## Discussion

Our main result in this paper is the completion of a portfolio of tools for the estimation of epidemiological indices from data. Such indices generalize current descriptions of infectious disease geographies by embedding general spatial effects and the actual reactivity of disease demographies. As a result, we have obtained a general framework that allows us to revisit current standards in the characterisation of the instantaneous state of epidemic transmission. In particular, by relaxing the assumption of spatially implicit disease dynamics, we have found that one may observe significant differences in alternative evaluations of the state of an epidemic. Comparative analyses suggest that a ranking of specific control measures may emerge depending on the degree of heterogeneity of community composition and of the related mobility fluxes. We thus completed the generalization of popular data-driven epidemiological estimators by deriving spatially explicit epidemicity indices to include information about human mobility and spatially-driven exposure to contagion. Our collection of results shows that adding the effects of mobility and spatial connectivity, as opposed to simple spatially implicit frameworks, allows one to better understand the behaviour of an ongoing epidemic and the role of each node on the collective fate. The spatially connected framework that we introduced here also contains the definition of epidemic subset, defined as the ensemble of local communities achieving reactive conditions.

Our results applied to synthetic data show that both the global effective RN and the various epidemicity indices display a major dependence on the local population size, on mobility fluxes among interconnected communities, and on local RNs. The results suggest that the computation of the global RN is essential to understand the epidemiological behaviour of a given metapopulation. Indeed, under a specific set of conditions, having some local effective RNs above the unit threshold does not necessarily lead to a ℛ^*G*^ *>* 1. This implies that the metapopulation may converge to an asymptotically stable DFE despite the fact that some communities might be characterized by effective RN values pointing at local endemic behaviour. Our results also confirm the dependence of the global RNs on the local ones, as previously proposed in the literature [23, 42, 43]. Here we improve over previous approaches by providing numerical schemes that can be directly applied in discrete time, which is useful when epidemiological indicators need to be computed from surveillance data. The tools described here can be implemented within state-of-the-art computational frameworks, like those used for real-time estimation of effective reproduction numbers via renewal equations [7, 20]. Therefore, they do not require the implementation of the time-marching solution to a fully coupled system of ODEs, e.g., a SIR-like compartmental model. While the long-term characterization of the dynamics of the system, captured by the global effective RN being either above or below the unit threshold, strongly depends on the magnitude and distribution of the involved population, as well as on the strength of the mobility fluxes, we find that the reactivity analysis based on the *ℓ*^1^-norm is substantially less dependent on such factors. This has relevant consequences. In fact, in order to secure non-reactive conditions in the whole metapopulation, all local communities must also display non-reactive conditions, i.e., all local effective RNs should be below the reactivity threshold [28].

We suggest that performing reactivity analyses using multiple output transformations may help to shed light on different characteristics of transient epidemic growth (measured in terms of, e.g., the sheer increase in the total number of infected individuals or the prevalence of infections in different communities). Using different norms may shift the attention from average trends to rapidly increasing local outbreaks, both of which can be useful as short-term warnings of potentially mounting pressure on the healthcare system at different spatial scales. Combining different output transformations and norms can be quite informative too. As an example, by having 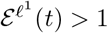 evaluated with the **Y**_**1**_ output and 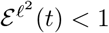 evaluated with the **Y**_**2**_ output signals that the potential burden on each local healthcare system may not increase in the short run, while the number of total infected individuals does.

The identification of the reactivity properties of an epidemiological system is thus essential to characterize carefully an epidemic outbreak. This is proven by our *ℓ*^1^-norm analysis on both the total number of infectious individuals or their prevalence in each local community. Under suitable conditions, the threshold that the spatially connected RN value should not exceed might be substantially lower than not only the unit—commonly seen as the divide between a hopeful or gloomy near future—but also the typically stricter spatially-implicit reactivity threshold [28]. However, when the mobility fluxes among communities are less marked, like in the case of our practical example on the covid-19 pandemic in Italy, this may not be the case, and the reactivity thresholds remain close to each other. This is caused by the similar populations and the comparable (and small) mobility fluxes across many Italian regions. Such thresholds would, therefore, be substantially different in the presence of higher mixing and heterogeneity among local populations.

Our key practical results are the global effective RN and the reactivity analysis in Italy. We find that non only the considered epidemicity indices always exceed the unit threshold, thereby suggesting reactive conditions throughout the period of analysis, but also all Italian regions were reactive during the considered timespan of the covid-19 epidemic in Italy. This observation has practical implications and justifies why, despite subthreshold estimates for the effective RNs during lull phases of the epidemic, notably during the 2020 and 2021 summers, local flare-ups have been so common throughout Italy. Furthermore, our correlation analyses showed that the considered epidemiological parameters are strongly associated with the largest local effective RN. This implies that, in order to secure non-reactive conditions, all local RNs should be smaller than a given threshold, which agrees with previous findings [28] and our claims stated hereinabove.

An overaching message of our analysis is that a spatial framework becomes essential, as opposed to simply useful and intellectually rewarding, in given circumstances characterized by a marked heterogeneity of population sizes, mobility fluxes, and local epidemiological conditions. For this reason, it is useful to retain the ability to mobilize these new spatially explicit tools when the level of heterogeneity in the disease geography, or boomimg demographic projections, imply forthcoming conditions conducive to major departures among the key indicators. In particular, this applies to epidemicity indices measuring the maximum growth rates of new infections. However elaborate, the spatially explicit version of the epidemicity index that we have derived here may provide results that sharpen decisively the prognostic worth of other epidemiological indicators based on reported infections. This is not only evident for airborne pathogens, such as the virus causing covid-19, but acquires a great importance also for waterborne diseases and for pathogens strongly depending on the environment, where heterogeneous local conditions must be considered to correctly describe the course of an ongoing epidemic [30].

We conclude that a portfolio of indicators based on epidemiological data (effective global RN and epidemicity indices tackling the reactivity of the system in both their spatially implicit and explicit versions) may constitute the best guarantee of a comprehensive overview of the effectiveness of epidemic controls under the typical uncertainty of real-time epidemic management conditions.

## Data availability statement

All codes and data involved in this study are available at the following public GitHub repository: https://github.com/cristianotrevisin/epidemiological-indices-metapopulation.

## Declaration

All authors declare no competing interests.

## Authors contributions

Conceptualization: CT, LM, MG, AR; Data Curation: CT, LM; Formal Analysis: CT, LM, MG, AR; Funding Acquisition: AR; Investigation: CT, LM, MG, AR; Methodology: CT, LM, MG, AR; Software: CT; Validation: CT, LM, MG, AR; Visualization: CT, MG; Writing – Original Draft Preparation: CT, AR; Writing – Review & Editing: CT, LM, MG, AR.

## Acknowledgement

C.T., and A.R. acknowledge funding from the Swiss National Science Foundation via the project ‘Optimal control of intervention strategies for waterborne disease epidemics’ (grant number 200021-172578). L.M. acknowledges funding from the Italian Ministry of University and Research through the project ‘Epidemiological data assimilation and optimal control for short-term forecasting and emergency management of COVID-19 in Italy’ (FISR 2020IP 04249).

